# Coordinated Cortical Thickness Alterations across Psychiatric Conditions: A Transdiagnostic ENIGMA Study

**DOI:** 10.1101/2022.02.03.22270326

**Authors:** MD Hettwer, S Larivière, BY Park, OA van den Heuvel, L Schmaal, OA Andreassen, CRK Ching, M Hoogman, J Buitelaar, DJ Veltman, DJ Stein, B Franke, TGM van Erp, ENIGMA ADHD Working Group, ENIGMA Autism Working Group, ENIGMA Bipolar Disorder Working Group, ENIGMA Major Depression Working Group, ENIGMA OCD Working Group, ENIGMA Schizophrenia Working Group, N Jahanshad, PM Thompson, SI Thomopoulos, RAI Bethlehem, BC Bernhardt, SB Eickhoff, SL Valk

**Affiliations:** Max Planck School of Cognition, Max Planck Institute of Human Cognitive and Brain Sciences, Leipzig, Germany; Institute of Neuroscience and Medicine, Brain & Behavior (INM-7), Research Centre Jülich, Jülich, Germany; Max Planck Institute for Cognitive and Brain Sciences, Leipzig, Germany; Institute of Systems Neuroscience, Medical Faculty, Heinrich Heine University Düsseldorf, Düsseldorf, Germany; Multimodal Imaging and Connectome Analysis Lab, McConnell Brain Imaging Centre, Montreal Neurological Institute and Hospital, McGill University, Montreal, Quebec, Canada; Department of Data Science, Inha University, Incheon, Republic of Korea; Center for Neuroscience Imaging Research, Institute for Basic Science, Suwon, Republic of Korea; Amsterdam UMC, Vrije Universiteit Amsterdam, Department of Psychiatry, Department of Anatomy & Neuroscience, Amsterdam Neuroscience, Amsterdam, The Netherlands; Centre for Youth Mental Health, The University of Melbourne, Parkville, Australia; Orygen, Parkville, Australia; NORMENT Centre, Division of Mental Health and Addiction, University of Oslo and Oslo University Hospital, Oslo, Norway; Imaging Genetics Center, Mark & Mary Stevens Neuroimaging and Informatics Institute, Keck School of Medicine, University of Southern California, Marina del Rey, CA, USA; Departments of Psychiatry and Human Genetics, Donders Institute for Brain, Cognition and Behaviour, Radboud University Medical Center, Nijmegen, The Netherlands; South African Medical Research Council Unit on Risk & Resilience in Mental Disorders, Dept of Psychiatry & Neuroscience Institute, University of Cape Town; Autism Research Centre, Department of Psychiatry, University of Cambridge, Cambridge, UK; Brain Mapping Unit, Department of Psychiatry, University of Cambridge, Cambridge, UK; Clinical Translational Neuroscience Laboratory, Department of Psychiatry and Human Behavior, University of California Irvine, Irvine Hall, room 109, Irvine, CA, 92697-3950, USA; Center for the Neurobiology of Learning and Memory, University of California Irvine, 309 Qureshey Research Lab, Irvine, CA, 92697, USA

**Author notes:** Correspondence to Meike D. Hettwer and Sofie L. Valk, Institute of Neuroscience and Medicine (INM-7: Brain and Behavior), Research Centre Jülich, Jülich, Germany; Institute of Systems Neuroscience, Medical Faculty, Heinrich Heine University Düsseldorf, Düsseldorf, Germany; Otto Hahn Group Cognitive Neurogenetics, Max Planck Institute for Human Cognitive and Brain Sciences, Leipzig, Germany.

**Keywords:** ENIGMA, psychopathology, magnetic resonance imaging, structural covariance, transdiagnostic

## Abstract

**Introduction:** Mental disorders are increasingly conceptualized as overlapping spectra with underlying polygenicity, neurodevelopmental etiology, and clinical comorbidity. They share multi-level neurobiological alterations, including network-like brain structural alterations. However, whether alteration patterns covary across mental disorders in a biologically meaningful way is currently unknown.

**Methods:** We accessed summary statistics on cortical thickness alterations from 12,024 patients with six mental disorders and 18,969 controls from the Enhancing NeuroImaging Genetics through Meta-Analysis (ENIGMA) consortium. First, we studied cortical thickness co-alteration networks as a form of pathological structural covariance. We identified regions exhibiting high inter-regional covariance across disorders (‘hubs’), and regions that strongly connect to these hubs facilitating network spreading of disease effects (‘epicenters’). Next, we applied manifold learning to reveal organizational gradients guiding transdiagnostic patterns of illness effects. Last, we tested whether these gradients capture differential cortical susceptibility with respect to normative cortical thickness covariance, cytoarchitectonic, transcriptomic, and meta-analytical task-based profiles.

**Results:** Co-alteration network hubs were linked to normative connectome hubs and anchored to prefrontal and temporal disease epicenters. The principal gradient derived from manifold learning captured maximally different embedding of prefrontal and temporal epicenters within co-alteration networks, followed a normative cortical thickness gradient, and established a transcriptomic link to cortico-cerebello-thalamic circuits. Moreover, gradients segregated functional networks involved in basic sensory, attentional/perceptual, and domain-general cognitive processes, and distinguished between regional cytoarchitectonic profiles.

**Conclusion:** Together, our findings indicate that disease impact occurs in a synchronized fashion and along multiple levels of hierarchical cortical organization. Such axes can help to disentangle the different neurobiological pathways underlying mental illness.

## INTRODUCTION

The conceptualization of mental disorders has undergone several transformations towards overlapping spectra of psychopathology (1,2) associated with underlying polygenicity, neurodevelopmental etiology, and epidemiological comorbidity (1,3,4). Efforts to empirically understand their dimensional structure has linked the general liability for mental illness to shared risk factors and common alterations in neurodevelopmental processes, predisposing to the clinical conditions ultimately manifested (5–8). Coordinated multi-level brain alterations across disorders may explain these phenomenological overlaps and common etiology.

Big-data neuroscience initiatives such as the Enhancing NeuroImaging Genetics through Meta-Analysis (ENIGMA) consortium have facilitated large-scale transdiagnostic investigations to identify shared and disorder-specific brain alterations (9). These studies consistently report cortical thickness alterations in mental illness (10–15), which serves as a proxy measure for neuronal density, cytoarchitecture, and intracortical myelination (16–18). Crucially, previous ENIGMA findings suggest that regional morphological alterations are not only shared between disorders (19–22), but also in part associated with shared genetic etiology (20), regional pyramidal-cell gene expression (21), microstructure and neurotransmitter system organization (22). While these findings highlight regional overlaps as shared effects between disorders, the current study aims to address inter-regional dependencies capturing coordinated transdiagnostic patterns of illness effects. That is, differences in brain morphology and function observed in psychiatric patients appear to follow network-like patterns constricted by underlying connectome organization (23–25). According to the nodal stress hypothesis, highly interconnected regions (‘hubs’) show higher susceptibility to pathological processes due to shared metabolic alterations, spread of pathogens, or similar gene expression profiles (25,26). In addition, regional disruptions can act as ‘disease epicenters’ by promoting pathological processes in areas they connect to, thus constituting anchors of network-like alterations (27). Although the role of network characteristics for cortical alterations in psychopathology is well established (25,28,29), it remains unknown how cross-disorder morphological alterations are embedded in a joint co-alteration network, and whether organizational principles shaping such a network link to underlying neurobiology.

An intuitive approach capturing inter-regional dependencies of illness effects is structural covariance of cortical thickness alterations, which forms cortex-wide co-alteration networks. While structural covariance partly reflects synchronized and genetically coupled maturation during healthy neurodevelopment (30–32), consolidated atrophy in illness also occurs more frequently in regions with high structural covariance (33,34). Moreover, inter-regional similarities in cortical features tend to be hierarchically organized: Previous mappings of low-dimensional cortex-wide gradients have described continua of cytoarchitectural complexity, long-versus short-distance connectivity, cell density, transcriptomic expression, and phylogenetic and ontogenetic timing (35–38). Such gradients (or ‘axes’) compactly summarize covariance patterns via connectome decomposition techniques (35,39), and place brain regions with similar covariance profiles closer together in a common coordinate-frame, regardless of their position on the cortex. These axes offer insights into the global arrangements of cortical features and appear to be distorted in several neuropsychiatric conditions (22,40–42). While the convergence of hierarchical neurobiological profiles thus appears to be a central feature of healthy brain organization, it is currently unknown whether the global arrangement of regional vulnerability to mental illness follows a hierarchical organization as well.

In this study, we identified hubs of transdiagnostic co-alteration networks and disease epicenters using meta-analytical maps for six mental disorders (autism spectrum disorder (ASD), attention-deficit/hyperactivity disorder (ADHD), major depressive disorder (MDD), schizophrenia (SCZ), bipolar disorder (BD), and obsessive-compulsive disorder (OCD)), provided by the ENIGMA consortium (10–15). We further employed a cortex-wide gradient mapping approach to identify hierarchical cortical arrangements of transdiagnostic illness effects. Last, we contextualized derived gradients with cytoarchitectonic and functional cortical profiles for multi-level evaluation. We performed multiple robustness checks to evaluate the stability of our findings.

## RESULTS

### Transdiagnostic covariance hubs inform disease epicenters

To study coordinated transdiagnostic effects of illness on cortical thickness, we accessed summary statistics from 12,024 patients with six mental disorders - ASD (10), ADHD (11), MDD (12), SCZ (13), BD (14), and OCD (15) – and 18,969 unaffected individuals from previously published ENIGMA studies (see Supplementary **Table S1**). Analyses were restricted to adult samples, except for ASD for which available summary statistics included all age groups. See Supplementary **Table S2** for information on sample demographics. For each condition, we retrieved a Cohen’s *d* map via the ENIGMA Toolbox (43) reflecting case-control differences in cortical thickness for 68 Desikan-Killiany parcels (44) (**Fig. 1A**). Cohen’s *d* maps were corrected for different combinations of covariates including age, sex, site and intelligence quotient (**Table S2**). For contextualization with normative network properties, we further accessed healthy control cortico-cortical structural (diffusion-weighted tractography; DTI) and functional (resting-state functional magnetic resonance imaging; rs-fMRI) connectivity data from an independent sample of healthy young adults from the Human Connectome Project (HCP; (45)) through the ENIGMA Toolbox (43) (**Fig. 1B**).

**Figure 1.**
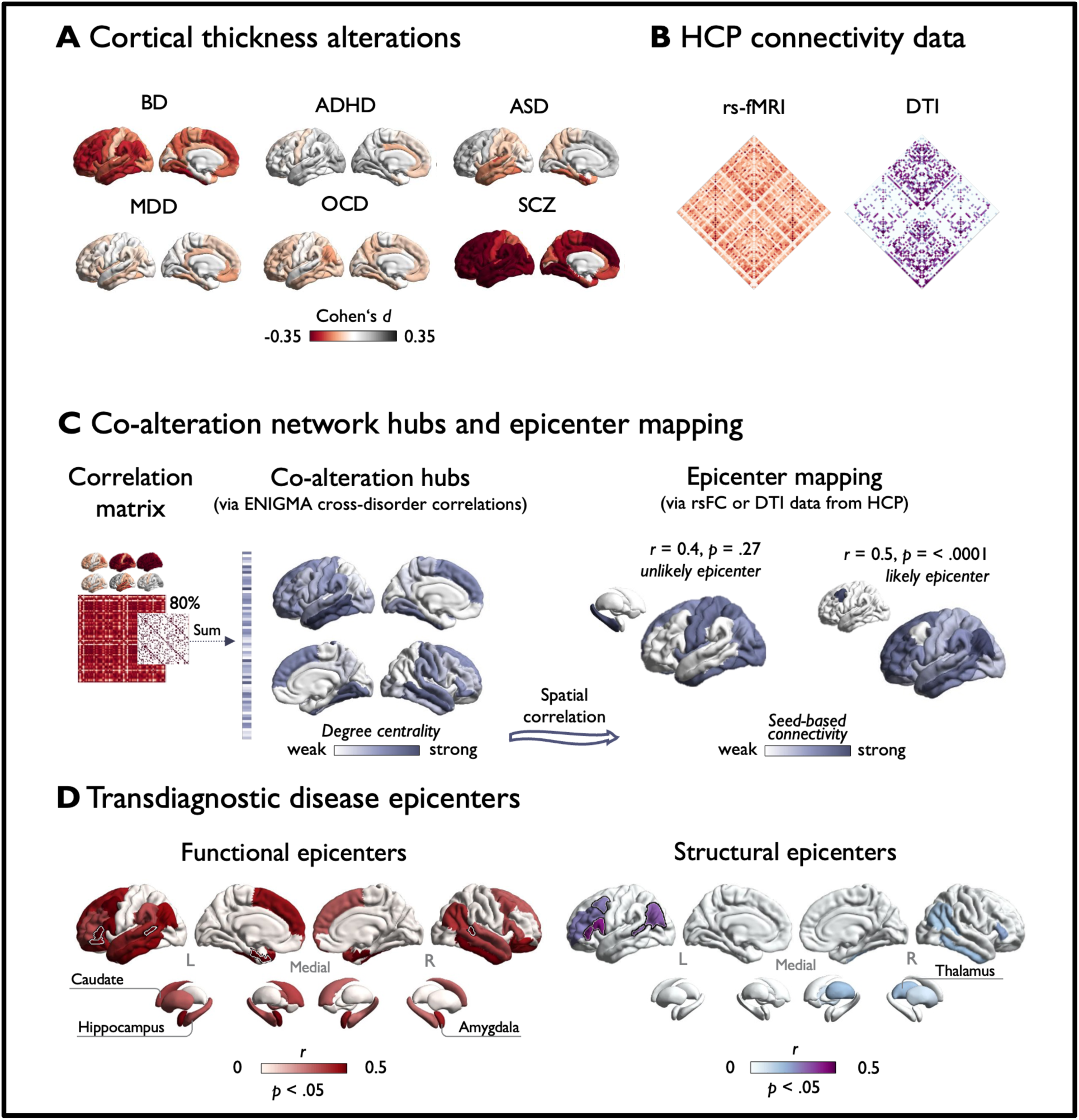
Hubs and epicenters shaping transdiagnostic co-alteration patterns. **A)** Condition-specific Cohen’s *d* maps indicating case-control differences in cortical thickness. **B)** Normative resting-state (rs-fMRI) and diffusion-weighted connectivity matrices from the Human Connectome Project (HCP; (45)**). C**) Left: Computation of co-alteration hubs. Degree centrality was computed as the sum of above-threshold (80%) connections at each parcel. Right: Visualization of the epicenter mapping approach. Seed-based connectivity profiles were systematically correlated with co-alteration hubs. **D)** Transdiagnostic disease epicenters are depicted as correlations between co-alteration hubs and normative seed-based connectivity profiles (rs-fMRI or diffusion tensor imaging (DTI)) from the Human Connectome Project (HCP), thresholded at p_spin_ < .05 (this figure shows DTI examples). High correlations imply high likelihood of a structure constituting a disease epicenter. Top five functional and structural disease epicenters are framed in white/black. ADHD = Attention-deficit/hyperactivity disorder; ASD = Autism spectrum disorder; BD = Bipolar disorder; MDD = Major depressive disorder; OCD = Obsessive-compulsive disorder; SCZ = Schizophrenia.

First, we computed a transdiagnostic co-alteration matrix by correlating Cohen’s *d* values between regions and across disorders. Regions showing a high sum of strong connections (i.e., correlations) were identified as co-alteration hubs (**Fig. 1C**). Transdiagnostic hub regions predominated in bilateral medial temporal gyrus and ventral temporal cortex, and more widespread in temporal and frontal regions. Findings were comparable at different thresholds (see **Fig. S1**). The spatial pattern of co-alteration hubs correlated with normative functional connectivity hubs (*r* = 0.50, p_spin_ < 0.0001), but less so with structural hubs (*r* = 0.18, p_spin_ = 0.08).

Having confirmed a general convergence between hubs of coordinated cortical thickness alterations and normative connectome organization, we next investigated whether these patterns are anchored to disease epicenters. Regions were identified as potential epicenters when their normative (HCP) seed-based cortex-wide connectivity profile correlated with the generated pathological co-alteration hub map (27).

Thus, the epicenter mapping approach highlights the role of regions that do not necessarily constitute hubs themselves (46) but may contribute to shaping the whole-brain manifestation of illness effects through strong or distributed connections with co-alteration hubs. Systematically investigating 68 cortical seeds revealed primarily temporal and prefrontal regions as potential transdiagnostic disease epicenters (**Fig. 1D**). Highest ranked functional disease epicenters were observed in the left entorhinal cortex, left *pars orbitalis*, right banks of the superior temporal sulcus (STS), left *pars triangularis*, and left STS (*r* = 0.55 to 0.59; all p_spin_ < .05). Top five structural disease epicenters were present in left pars opercularis and triangularis, inferior parietal lobe, STS bank, and caudal middle frontal gyrus (*r* = 0.28 to 0.42; all p_spin_ < .05).

### Macroscale gradients of transdiagnostic co-alteration networks

So far, our analyses suggest that the cortex-wide network of transdiagnostic illness effects is non-randomly organized, with hubs of prominent covariance and epicenters shaping the co-alteration network. Next, via manifold learning, we sought to study the embedding of these features within low-dimensional organizational gradients (39,47). This analysis was based on the same co-alteration matrix used to derive transdiagnostic hubs (**Fig. 1C** & **2A**). We applied diffusion map embedding (47) to project regional and long-range connections within covariance networks into a common space. This yielded unitless components, each of which denotes the position of nodes on a continuum describing similarities in regions’ structural covariance profiles (**Fig. 2A**). Thus, opposing apices of a gradient reflect maximally divergent covariance patterns.

**Figure 2.**
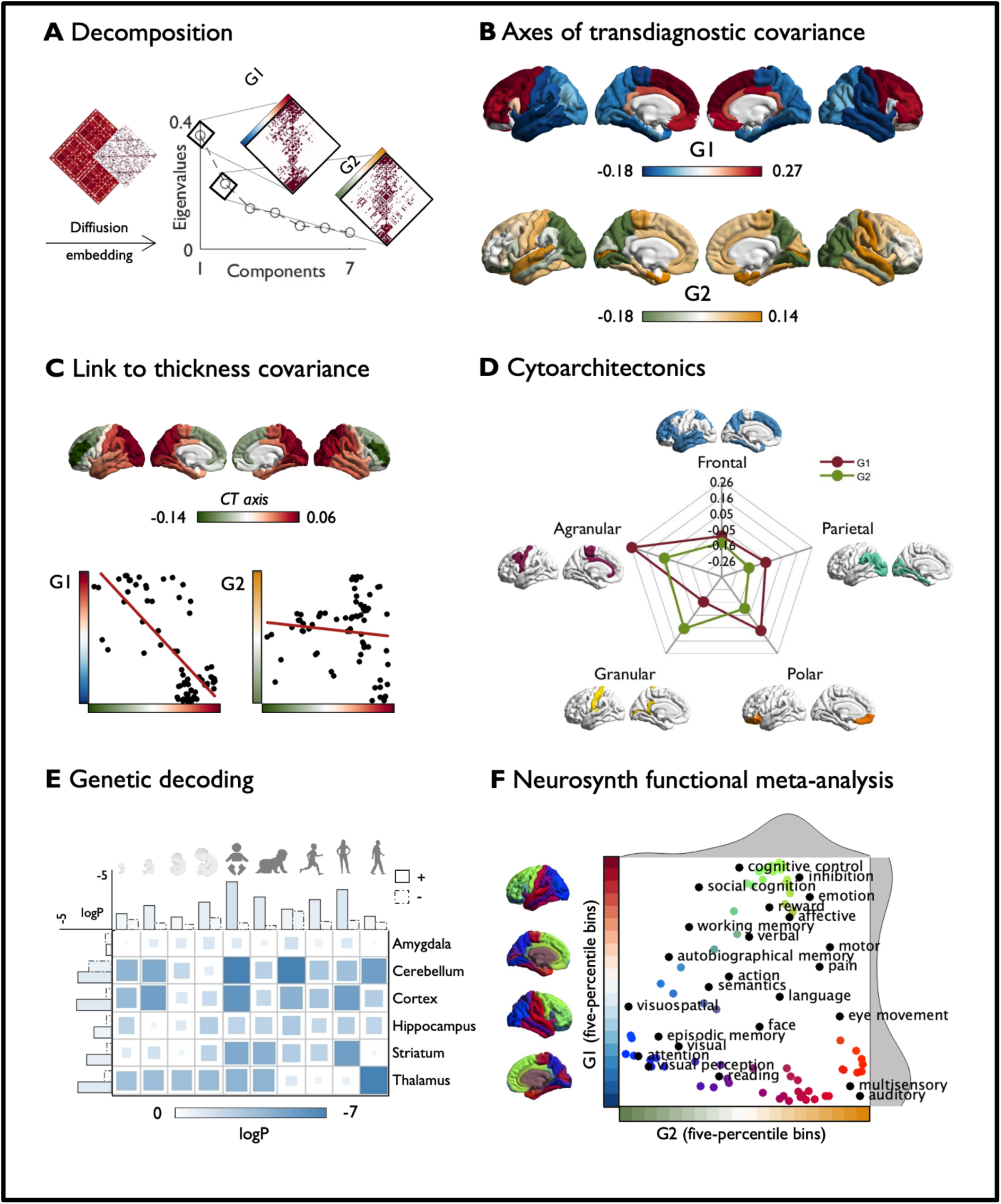
Macroscale organization of transdiagnostic covariance in cortical thickness alterations. **A)** A cross-condition structural covariance matrix was thresholded at 80% and decomposed using diffusion map embedding. Covariance along the principal (G1) and second (G2) gradients is depicted on the right. **B)** Transdiagnostic gradients G1 and G2. **C)** Correlation between a normative axis of cortical thickness covariance (36) and transdiagnostic gradients. **D)** Cross-condition gradients stratified according to von Economo-Koskinas cytoarchitectonic classes (49). **E)** Gene expression across brain structures and developmental phases based on genes for which spatial expression patterns correlated with G1. The matrix depicts average results for genes that spatially correlated positively (i.e., more strongly expressed in prefrontal apex) or negatively (i.e., more strongly expressed in temporal apex) with G1. Bars differentiate between positively (*solid edges*) and negatively (*dashed edges*) correlated genes. **F)** Meta-analysis for diverse cognitive functions obtained from NeuroSynth (56) similar to Margulies et al. (35). We computed parcel-wise *z*-statistics, capturing node-function associations, and calculated the center of gravity of each function along 20 five-percentile bins of G1 and G2. Function terms are ordered by the weighted mean of their location along the gradients.

The principal gradient of transdiagnostic covariance (G1) captured a dominant dissociation between frontal and temporal lobes and accounted for 36% of variance in transdiagnostic co-alteration (**Fig. 2B**). G1 correlated with spatial cortical thickness alteration maps of ADHD, ASD, and OCD (**Table S3**). The secondary gradient (G2) spanned from occipito-parietal regions to temporo-limbic structures, explaining 21% of variance. G2 correlated significantly with spatial cortical thickness alteration maps of SCZ, BD, ADHD, ASD, and OCD. Findings were comparable at different thresholds and robust against parameter manipulation (see **Fig. S2** and **S3**) and selection of diagnoses (see **Fig. S4**). An overview of all computed gradients is presented in **Fig. S5**.

Investigating the correspondence between the disease epicenters and the gradient-based cross-condition maps, we found that the apices of G1 captured previously identified functional disease epicenters (**Fig. S6**). This implies that frontal and temporal epicenters each contribute to the overall pattern of co-alterations but do so in a distinct manner (**Fig. S7**).

Since previous studies have shown that cortical thickness alterations in psychopathology are more prominent in regions with high structural covariance (48), we assessed whether the disease-related relative changes in cortical thickness align with normative organization of absolute cortical thickness. Indeed, we observed a correlation between the principal cortical thickness covariance gradient (anterior-posterior; **Fig. 2C**) (36) and G1 (*r* = -0.74, p_spin_ = .0015) but not G2 (*r* = -0.11, p_spin_ = .27). The second cortical thickness covariance gradient (inferior-superior) was not related to G1 (*r* = 0.32, p_spin_ = 0.21) or G2 (*r* = -0.25, p_spin_ = .07).

### Microstructural and transcriptomic contextualization

After capturing macroscale organization of disease effects, we contextualized identified gradients with microscale cytoarchitecture to gain a multi-level understanding of neurobiological cortical profiles in shaping transdiagnostic co-alteration networks. To this end, we stratified our gradients according to von Economo-Koskinas cytoarchitectonic classes (49). We observed a prominent distinction between granular and agranular cortices across our principal transdiagnostic gradient (**Fig. 2D**), whereas G2 distinguished between granular and parietal cytoarchitectonic classes.

Using *post-mortem* gene expression data from the Allen Human Brain Atlas (AHBA) as a reference (50), we next identified genes for which spatial expression patterns significantly correlated with G1 (FDR p < .001; see **Table S4**). This approach has previously revealed genetic links to normative brain development and organization (50–52) as well as structural abnormalities in disease (42,53,54). Out of 199 genes for which expression patterns correlated significantly with G1, 136 showed a positive correlation with G1, meaning that they were more strongly expressed in the PFC than in temporal regions. Developmental gene enrichment analyses (55) revealed that, next to the cortex, identified genes were most prominently expressed in the cerebellum and thalamus across various developmental windows (**Fig**. 2E). In a combined assessment of all brain structures, genes appeared to be enriched most strongly during neonatal early infancy, early childhood, and adolescence. G2 was not significantly associated with genes included in the AHBA.

### Associations with task-based functional activations

Lastly, we aimed to identify potential functional implications by investigating whether organizational axes dissociate between task-specific functional activations. To this end, we conducted a meta-analysis on 24 cognitive terms using the NeuroSynth database (56). We defined regions of interest as five-percentile bins of both gradients and studied the distribution of functional networks along the axes via *z*-statistics. This analysis revealed that combined features of both gradients distinguish between different co-alteration patterns in primary (e.g., ‘auditory’) and ‘multisensory’ regions at the temporal apex, higher-order perceptual structures (e.g., ‘visual perception’ and ‘attention’) at the occipito-parietal apex and complex cognitive functions (e.g. ‘cognitive control’ and ‘inhibition’) at the frontal apex (**Fig**. 2F).

## DISCUSSION

Our study reports coordinated effects of six major mental disorders (SCZ, BD, OCD, ASD, ADHD, and MDD) on cortical thickness and their association with functionally relevant neurobiological patterns across multiple scales of analysis. Thus, we extended previous investigations of shared regional effects (19–22) towards a network-based approach that embeds regional alterations within cortical hierarchies of transdiagnostic covariance of illness effects.

We identified hubs of transdiagnostic co-alteration predominantly in lateral and ventral temporal lobes, with some impact on post-central and medial frontal regions. As they followed the spatial pattern of normative functional connectivity hubs, captured susceptibility may likely link to nodal stress (25,26,57). Indeed, *in vivo* markers of e.g. aberrant energy metabolism and *post-mortem* proteomic analyses have revealed overlap between MDD, SCZ, and BD (58–60). As hubs are more strongly influenced by genes than non-hubs (61), hub regions may exhibit increased shared vulnerability for atypical neurodevelopment, as seen in polygenic and genetically overlapping psychiatric diagnoses. Thus, nodal stress, along with other potential factors such as shared genetic susceptibility, appears to be a strong candidate explanation for the irregular topographic distribution of covarying illness effects (28,62).

Present results further indicate that large-scale patterns of shared illness effects are shaped by both structural and functional epicenters, where influences of functional epicenters emerge above and beyond hard-wired tracts. A certain divergence between structural and functional patterns is likely (63), as functional connectivity reflects a temporal correlation of activity which may be driven by distant input into a spatially distributed polysynaptic network (64,65). Here, we observed highest concordance with co-alteration hubs in connectivity profiles of prefrontal and temporal regions. Thus, epicenters preferentially emerged in regions known to extend long-range connections (66), facilitating their contribution to cortex-wide organizational patterns. Structures in the mediotemporal lobe and ventrolateral PFC were identified as most likely epicenters. Both regions have been implicated in cognitive impairments and developmental susceptibility across mental disorders (51,67,68). More generally, mediotemporal structures act as nodal points between multimodal cortical association areas and the subcortex, and feature transitions in cytoarchitecture from iso- to allocortical regions (37). These features may increase both vulnerability to nodal stress and the spread of pathological alterations through wide-ranging connections. The vlPFC shows protracted plasticity throughout multiple neurodevelopmental stages (69). While allowing for continuous refinement of complex cognitive abilities, protracted plasticity gives room for aberrant maturational processes leaving the individual more susceptible to developmental aberrations. Overall, the epicenter mapping approach thus identified anchors of large-scale transdiagnostic co-alteration networks in regions that both have the potential to spread illness effects through long-range connections and are susceptible to maturational aberration.

Further investigating transdiagnostic covariance via manifold learning, we recapitulated cortex-wide gradients along which co-alteration patterns were organized. The principal transdiagnostic gradient captured a cortex-wide segregation of frontal and temporal structures, indicating that cortical thickness alterations in both regions are embedded in maximally distinct covariance networks. The concordance of G1 with the normative organizational axis of cortical thickness covariance (36) mirrors previous findings indicating increased susceptibility to cortical atrophy in regions that exert high structural covariance (33,70). As cortical thickness covariance is assumed to reflect common maturational trajectories (30), atypical neurodevelopment likely contributes to shaping cortical gradients of co-alteration networks. The process of shaping transdiagnostic gradients throughout development may further be influenced by subcortico-cortical circuits (71–74), as suggested by our transcriptomic decoding findings. Here, we observed that genes whose expression pattern align with G1 are also enriched in the cerebellum and thalamus in early developmental phases. Notably, studies on subcortical interactions have linked impaired functional coordination within cerebello-thalamo-cortical circuits to a general liability for psychopathology (75,76). It is thus possible that the organization of transdiagnostic co-alterations observed in the cortex partly builds upon alterations in subcortical circuits. The degree to which these alterations are shared between subcortical and cortical structures, and the transdiagnostic and disease-specific value of such interrelations may be investigated in future studies. The secondary gradient was restricted to uni- and heteromodal sensory cortices in the posterior cortex, segregating regions that hold primary sensory (pericalcarine cortex, post-central and superior temporal gyrus) and paralimbic (entorhinal) cortices from multimodal association regions in the occipito-parietal cortex. Both axes described segregation along different cytoarchitectural classes. Whereas G1 traversed between agranular, paralimbic, versus granular, primary cortices, G2 showed a cytoarchitectural divergence between granular and frontal/parietal cortices. Variable susceptibility to disease impact thus suggests that areas with shared cytoarchitecture are more likely embedded similarly within pathological networks. This may be due to similar local computational strategies supported by cell count and wiring strategies (17), development (77), and the degree of plasticity associated with different degrees of cortical lamination (51). Future work may further investigate the specific neurobiological mechanism linking cytoarchitecture, function, and mental illness.

Lastly, we contextualized our findings with respect to functional processes through meta-analytical task-based activations. Combining G1 and G2 in a two-dimensional space revealed distinct co-alteration profiles at three levels of information processing, i.e. primary/multi-sensory, perception/attention, and domain-general cognitive control. Interestingly, all three levels show various processing impairments in different neuropsychiatric conditions which are in part interrelated: 1) Atypical early development of sensory cortices can contribute to social cognitive deficits through impaired social cue perception (78,79) and, more generally, deficits in multisensory binding (79,80). 2) At an intermediary level, aberrant functional involvement and structural integrity of attention networks have been identified as a prominent transdiagnostic feature of neuropsychiatric conditions (28,81). 3) Upstream consequences of dysregulated attention networks ultimately contribute to impaired higher-order cognitive functions such as executive control. Impaired executive control does not only constitute a transdiagnostic feature in mental illness (82), but is also a predictor of cognitive and socio-occupational impairment (82–84). Despite inter-related deficits within functional networks, the fact that multiple processing levels are associated with distinct structural co-alteration patterns indicates independent maturational causes and distinct vulnerability. In line with findings from cytoarchitectonic contextualization, levels of functional engagement of cortices involved in similar tasks appear to leave brain regions processing similar types of information with shared susceptibility. Given that sensory regions develop earlier than association regions in the cortical maturational sequence (85), differences in pathological covariance profiles may link to the degree to which their developmental peaks overlap with vulnerable periods for mental disorders. Together, these findings raise the question whether identified cortical gradients also reflect a spatio*temporal* gradient of atypical neurodevelopment and inspire respective investigations in longitudinal/prospective studies.

Though our findings underline the relevance of transdiagnostic approaches, they do not contradict the existence of etiological and phenomenological differences between psychiatric diagnoses. Rather, hierarchical transdiagnostic gradients shall complement disorder-specific insights, and individual disorders appear to vary in their cortical profiles along the two identified cortical axes. Although we mostly included adult samples and age-corrected summary statistics, there are some offsets among mean ages of ENIGMA maps and between ENIGMA maps and the reference data from other sources (e.g. HCP), which potentially influenced parameters known to change during development such as hub organization (85). Additionally, neurodevelopmental conditions have different mean ages of onset so that patients included have certainly experienced different lengths of disease and medication histories. It should further be noted that current findings may not be generalizable to neuropsychiatric conditions not included in our study. Lastly, ENIGMA summary statistics used here are based on the Desikan-Killiany atlas (44). They thus contain comparatively sparse data points across the cortex and summarize data from broader areas that contain a mosaic of neurobiological regions that may be differentially affected by disease. Moreover, differences in parcel size (86), measurement error, subject motion and scanner/site effects (87,88) may slightly influence spatial covariance analyses.

In sum, our findings highlight the value of linking multiple neurobiological levels of information - from macroscale neuroimaging to microscale transcriptomic data - to identify systematic transdiagnostic patterns of illness effects. Investigating these patterns revealed coordinated cortical alterations across conditions that are shaped by connectomic, cytoarchitectonic, and functional characteristics. Future work will expand on this approach not only to include different modalities and neuroimaging metrics (e.g. surface area and subcortical structures), but also to consider a much wider range of conditions and age ranges, which is now becoming increasingly possible due to the availability of multi-disease consortia and datasets (89,90). This provides a crucial step towards understanding the neuro-etiology of neuropsychiatric conditions.

## METHODS

### ENIGMA Neuroimaging summary statistics

For our transdiagnostic analyses, we used publicly available multi-site summary statistics published by the ENIGMA Consortium, and available within the ENIGMA Toolbox (https://github.com/MICA-MNI/ENIGMA; (43)). Included mental disorders comprised ADHD (11), ASD (10), BD (type I and II, cumulated) (14), MDD (12), OCD (15), and SCZ (13). Except for ASD for which available summary statistics included all age groups, we restricted our analyses to adult samples. This decision may increase the variance in disease duration due to differences in typical ages of onset associated with the six diagnoses, however, we aimed to match adults to minimize effects that are linked to development and aging, which are potentially larger than the effects of disease duration. We based our analyses on covariate-adjusted case-control differences denoted by across-site random-effects meta-analyses of Cohen’s *d*-values for cortical thickness. Age, sex, and site information was fitted to cortical thickness measures via multiple linear regression analyses. See **Table S2** for an overview on demographics and study-specific covariates. Preceding the computation of summary statistics, raw data was pre-processed, segmented and parcellated according to the Desikan-Killiany atlas (44) in FreeSurfer (http://surfer.nmr.mgh.harvard.edu) at each site and according to standard ENIGMA quality control protocols (see http://enigma.ini.usc.edu/protocols/imaging-protocols). Sample sizes ranged from 1,272 (ADHD) to 9,572 (SCZ). We redirect the reader to the original publications (10–15) for more details on age matching and controlling for medication or comorbidities. Ethics approval and subjects’ informed consent was obtained by individual cohort investigators.

### Population connectivity data

Functional and structural connectivity matrices were based on 1 hour of rs-fMRI and diffusion MRI from healthy adults (*n* = 207, 83 males, mean age = 28.73 ± 3.73 years), respectively. The data was acquired through the HCP (45), minimally pre-processed according to HCP guidelines (91) and made publicly available as group-average structural and functional connectivity matrices in the ENIGMA Toolbox (43). The computation of connectivity matrices is described in more detail elsewhere (27) (and see Supplementary Material).

### Structural covariance of disease effects on local brain structure

We derived a 68×68 cross-condition correlation matrix by computing inter-regional Pearson’s correlations of cortical thickness Cohen’s *d* values across the six conditions.

### Covariance hubs and transdiagnostic disease epicenters

In order to derive co-alteration network hubs using a degree centrality approach, we first identified which connections (i.e., correlations) of the previously derived cross-condition correlation matrix belong to the top 20% of strong connections. We then computed the sum of these connections for each parcel, where regions with many strong connections represent hubs of high transdiagnostic covariance of illness effects (Fig. 2A). We then accessed whole-brain functional (rs-fMRI) and structural (DTI) connectivity matrices from a healthy adult HCP dataset (45) via the ENIGMA Toolbox (43), which we also thresholded at 80%. Normative connectivity hub maps based on HCP data was computed using the same degree centrality approach (i.e., the sum of all connections, thresholded) and spatially correlated with the transdiagnostic structural co-alteration hub map. Significance was assessed via spin tests (see Supplementary Material and (92)).This analysis aimed to assess whether co-alteration hub regions align with the normative underlying connectome and may thus be linked to nodal stress.

To identify transdiagnostic disease epicenters, we systematically assessed spatial similarity of each parcel’s normative whole-brain connectivity profile with our map of co-alteration hubs using spatial permutation tests. To do so, we collected seed-based functional (rs-fMRI) and structural (DTI) connectivity matrices for each parcel and 14 subcortical structures from the same HCP dataset (45). We then spatially correlated each structure’s connectivity profile with the co-alteration hub map. The higher the spatial similarity between an epicenter’s connectivity profile and co-alteration hubs, the more likely this structure represents a disease epicenter (at *p* < .05 after spin tests). Resulting likelihoods were ranked to identify the top five structural and functional transdiagnostic disease epicenters.

### Gradient decomposition

We computed macroscale organizational gradients using BrainSpace (https://github.com/MICA-MNI/BrainSpace; (47)) in Matlab 2020b. The 68×68 structural covariance matrix was thresholded at 80% and transformed into a non-negative square symmetric affinity matrix by using a normalized angle similarity kernel. We then applied diffusion mapping as a nonlinear dimensionality reduction method (39,47) to estimate the low-dimensional embedding of our previously derived high-dimensional affinity matrix. Here, cortical nodes that are close together reflect nodes that are inter-connected by either many supra-threshold or few very strong edges, whereas nodes that are farther apart reflect little or no covariance. We set α, a parameter which controls the impact of sampling density (where 0 to 1 = maximal to no influence), to 0.5. This α value retains global relations in the low-dimensional space and is assumed to be comparatively robust to noise in the input matrix. Lastly, we assessed the amount of information explained by received gradients, selected the first two gradients for further analyses and projected them onto a cortical mesh using BrainStat (https://github.com/MICA-MNI/BrainStat).

### Link to normative axes of cortical thickness organization

An association with normative cortical thickness organization was studied by correlating derived transdiagnostic gradients with previously established gradients of cortical thickness covariance in healthy adults. These two normative gradients were based on cortical thickness data from individuals in the S1200 HCP sample and were derived using the same diffusion embedding approach as described in this manuscript (see Valk et al. (36) for details). Spatial associations were evaluated using spin tests (92).

### Cytoarchitectonic contextualization

To determine whether transdiagnostic gradients recapitulate cytoarchitectonic variation evidenced by *post-mortem* histological assessments, we further stratified our gradients according to the five von Economo-Koskinas cytoarchitectonic classes (49). This atlas subdivides the cortex into five categories: agranular (thick cortex housing large cells but scarce layers II and IV), frontal (thick cortex, large but sparse cells, layers II and IV are present), parietal (thick cortex that is rich in cells, dense layers II and IV, slender pyramidal cells), polar (thin cortex, rich in cells, particularly granular cells) and granular/konicortex (very thin cortex with highest density of small cells).

### Genetic decoding

Having established macro- and microscale contextualization of our findings, we finally aimed to understand its association with gene transcriptomic data. Initially, we correlated our gradients with the *post-mortem* gene expression maps provided by the Allen Institute for Brain Science (AIBS) (50). Microarray expression data was processed in *abagen* (93), including intensity-based filtering, normalization and aggregation within Desikan-Killiany parcels and across donors. Only genes with a similarity of r > 0.2 across donors were included, resulting in 12,668 genes for the analysis (43). We thresholded the spatial correlation between microarray expression data and our gradients at FDR p < .001. Next, we fed identified significant genes into enrichment analysis via the cell-type specific expression analysis (CSEA) developmental expression tool (http://genetics.wustl.edu/jdlab/csea-tool-2) (55). This allowed us to compare genes identified with respect to the AIBS repository with developmental expression profiles from the BrainSpan dataset (http://www.brainspan.org), yielding more detailed, yet indirect, information about brain structures and developmental windows in which identified genes are enriched.

### Functional decoding

To assess whether transdiagnostic gradients capture differential impact on cognitive networks, we assessed the distribution of various cognitive functions along transdiagnostic gradients. To this end, we conducted a meta-analysis using the NeuroSynth (56) database as described by Margulies et al. (35). Briefly, we derived 20 region of interest maps from five-percentile bins of G1 and G2 and examined their association with 24 topic terms via z-statistics. Topic terms were then sorted based on their center of gravity and arranged in a two-dimensional space that was created by merging G1 and G2, for visualization.

## Data Availability

All data produced will be made available online at https://github.com/CNG-LAB/cngopen/transdiagnostic_gradients

## Acknowledgements

Many scientists around the world contributed to ENIGMA but did not take part in the writing of this report. A full list of contributors to ENIGMA is available at http://enigma.ini.usc.edu/about-2/consortium/members/. The authors would like to express their gratitude to the open science initiatives that made this work possible: (i) the ENIGMA Consortium (core funding for ENIGMA was provided by the NIH Big Data to Knowledge (BD2K) program under consortium grant U54 EB020403 to P.M.T.), (ii) The Allen Human Brain Atlas and the abagen toolbox (https://doi.org/10.5281/zenodo.4984124), and (iii) the Human Connectome Project (principal investigators David Van Essen and Kamil Ugurbil; U54 MH091657), funded by the 16 NIH institutes and centers that support the NIH Blueprint for Neuroscience Research and by the McDonnell Center for Systems Neuroscience at Washington University.

## Data availability

All data analyzed in this manuscript were obtained from open-access sources. Disorder-specific Cohen’s *d* maps derived from ENIGMA meta-analyses were accessed via the ENIGMA Toolbox (https://enigma-toolbox.readthedocs.io/en/latest/; (43)). Through the toolbox, we also accessed normative connectivity data from a Human Connectome Project young adult sample (HCP; http://www.humanconnectome.org/; (45)), the von Economo-Koskinas cytoarchitectonic atlas (49), and gene transcriptomic data from the Allen human brain atlas (https://human.brain-map.org/). Further genetic analyses were performed using the cell-specific enrichment analysis tool (http://genetics.wustl.edu/jdlab/csea-tool-2/). The functional meta-analysis was based on the NeuroSynth database (https://neurosynth.org/). Gradient mapping analyses were based on open-access tools (BrainSpace, https://brainspace.readthedocs.io/en/latest/). Visualizations were carried out using BrainStat (https://github.com/MICA-MNI/BrainStat) in combination with ColorBrewer (https://github.com/scottclowe/cbrewer2).

