## Supplementary Material for "Coordinated Cortical Thickness Alterations across Psychiatric Conditions: A Transdiagnostic ENIGMA Study"

### Supplementary Methods

*Population connectivity data*

Population connectivity data was derived from a healthy young adult sample (n=207; 83 males, mean age±SD=28.73±3.73 years, range=22-36 years) from the Human Connectome Project (HCP; (1)). Resting-state functional data underwent distortion and motion corrections, intensity inhomogeneity corrections and intensity normalization, brain extraction, normalization to MNI152 space and projection onto the cortical surface. Pre-processing of diffusion MRI data included b0 intensity normalization as well as corrections for head motion, susceptibility distortion and eddy currents. Both functional and structural connectivity data was parcellated according to the Desikan-Killiany atlas (2).

Subject-level functional connectivity matrices were generated by pair-wise correlations between time series of 68 cortical parcels and 12 sub-cortical structures. Z-scored subject-level data was accumulated to derive a group-average functional connectome (3). Structural connectivity matrices included in the ENIGMA Toolbox are based on anatomically constrained tractography, where reconstructed streamlines were generated for 68 parcels and 12 sub-cortical structures. Using distance-dependent thresholding, a group-average structural connectome was derived and log-transformed.

*Spin tests*

Wherever possible, we implemented spin tests to assess the significance of spatial similarities via permutations. Spatial permutation tests correct for auto-correlations between smooth spatial maps by generating null models of respective spatial overlaps. That is, coordinates of cortical data are inflated to a sphere and rotated 1000 times, matching phenotypic data to different parcels in every permutation (4). Significance is determined by testing initial correlation coefficients against the null distributions retrieved by correlating rotated spatial maps.

**Supplementary Results**

*Robustness of cross-disorder co-alteration network hubs*

In order to assess the stability of cross-disorder co-alteration hub maps, we recreated hub maps based on cross-disorder inter-regional correlation matrices thresholded at 90%, 70% and 50% (Figure S1). All three alternative hubs maps correlated significantly with original co-alteration hubs (90% threshold: $r = 0.78$; 70% threshold: $r = 0.91$; 50% threshold: $r = 0.62$; all $p_{spin} < .05$).

*Robustness of transdiagnostic gradients*

To assess robustness of the first two transdiagnostic gradients, we compared them to gradients derived from manipulating analysis steps in the gradient computation and changing parameters in the BrainSpace toolbox (5) (see Figure S2 and S3). First, since there were generalized differences in the strength of disease impact on cortical thickness across disorders, we mean-corrected the initial correlation matrix by using a partial correlation coefficient. Gradients computed based on this mean-corrected matrix correlated highly with original gradients (G1: $r = 0.94$; G2: $r = 0.87$). Second, original gradients correlated highly with gradients derived using a different non-linear dimension reduction method (Laplacian eigenmap: G1: $r = 1$; G2: $r = 0.99$) or a linear dimension reduction method (principal component analysis: G1: $r = 1$; G2: $r = 0.99$). Third, even though data was normally distributed, it yielded relatively sparse data points for each inter-regional correlation. We therefore tested whether the use of Spearman's rho instead of Pearson's $r$ in computing the initial correlation matrix influences gradient organization. Gradients based on Spearman's rho correlation coefficients correlated highly with the original gradients (Spearman: G1: $r = 0.94$; G2; $r = 0.84$). Last, original gradients also correlated highly with gradients for which cut-off values (i.e. sparsity) of the correlation matrix was manipulated (sparsity of 90%: G1: $r = 0.97$; G2: $r = 0.92$; sparsity of 70%: G1: $r = 0.99$; G2: $r = 0.96$; sparsity of 50%: G1: $r = 0.94$; G2: $r = 0.80$). Overall, original gradients were robust against a number of parameter manipulations.

*Influence of individual disorders on gradient organization*

To assess whether gradient organization was differentially impacted by individual disorders, we performed leave-one-disorder-out analyses and correlated resulting gradients with original gradients G1 and G2 (Figure S4). We observed that gradients were generally robust against leaving out single disorders (w/o ADHD: $r_{G1} = 0.95$; $r_{G2} = 0.77$; w/o BD: $r_{G1} = 0.99$; $r_{G2} = 0.86$;

w/o SCZ: $r_{G1} = 0.98$; $r_{G2} = 0.94$; w/o OCD: $r_{G1} = 0.99$; $r_{G2} = 0.83$; w/o MDD: $r_{G1} = 0.99$; $r_{G2} = 0.97$). However, this was not the case for ASD (without ASD: $r_{G1} = 0.10$; $r_{G2} = 0.01$). Leaving out ASD in gradient computations appeared to lead to a switch in features to be reflected in principal and secondary gradients, as was observed in significant correlations of the principal gradient without ASD with the original G2 ($r = 0.86$, $p_{spin} < .001$) and of the secondary gradient without ASD with the original G2 ($r = 0.48$, $p_{spin} < 0.01$). This finding is not surprising, as cortical thickness alterations in ASD show a spatial pattern that is highly similar to G1 and thus likely strengthens the weight of features then represented in G1.

*G1 captures segregation of functional disease epicenters*

As we noticed that frontal and temporal disease epicenters appear to be segregated by G1 (see Figure S6) suggesting differential impact on co-alteration network organization, we performed a follow up analysis to confirm this assumption. In order to evaluate whether whole-brain cross-disorder disease impact shows different covariance patterns for frontal and temporal epicenters, we A) extracted cross-disorder inter-regional correlations from the 68 x 68 correlation matrix (see Figure 2A & 3A of the main manuscript) for all frontal and temporal disease epicenters, respectively, and computed their degree centrality as the sum of all correlations of each epicenter parcel. B) We extracted cross-disorder whole-brain structural covariance for two representative epicenters, the left *pars orbitalis* and entorhinal cortex, which emerged as the two strongest (functional) epicenters. Both approaches revealed that frontal epicenters show covariance of disease impact across wide-spread regions of the cortex, whereas temporal epicenters show highest correlations within temporal, and no correlations with frontal regions (see Figure S7).

### Supplementary Figures

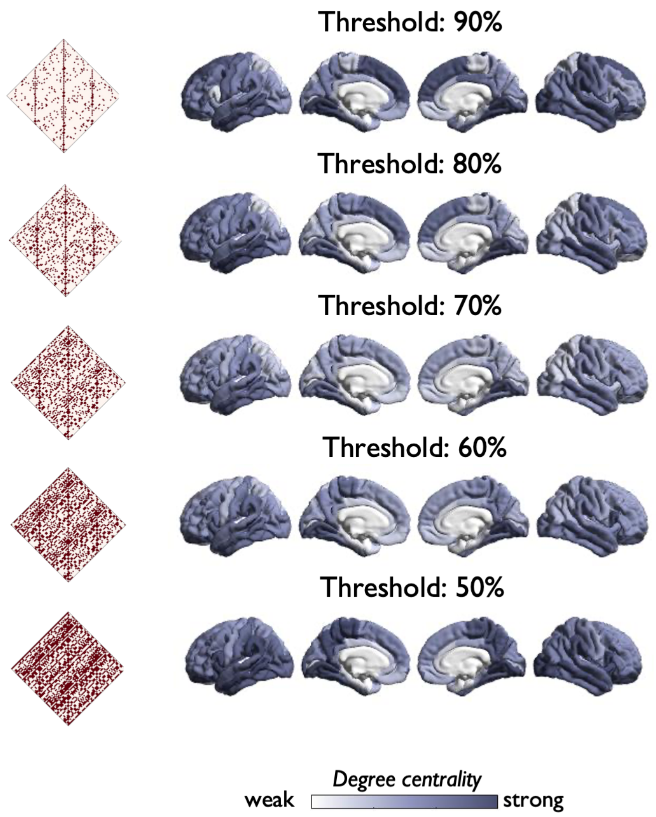


**Figure S1**. *Transdiagnostic co-alteration network hubs at different thresholds.* In order to test the stability of co-alteration hubs, we recreated hub maps based on correlation matrices (left) thresholded at 50 to 90%.

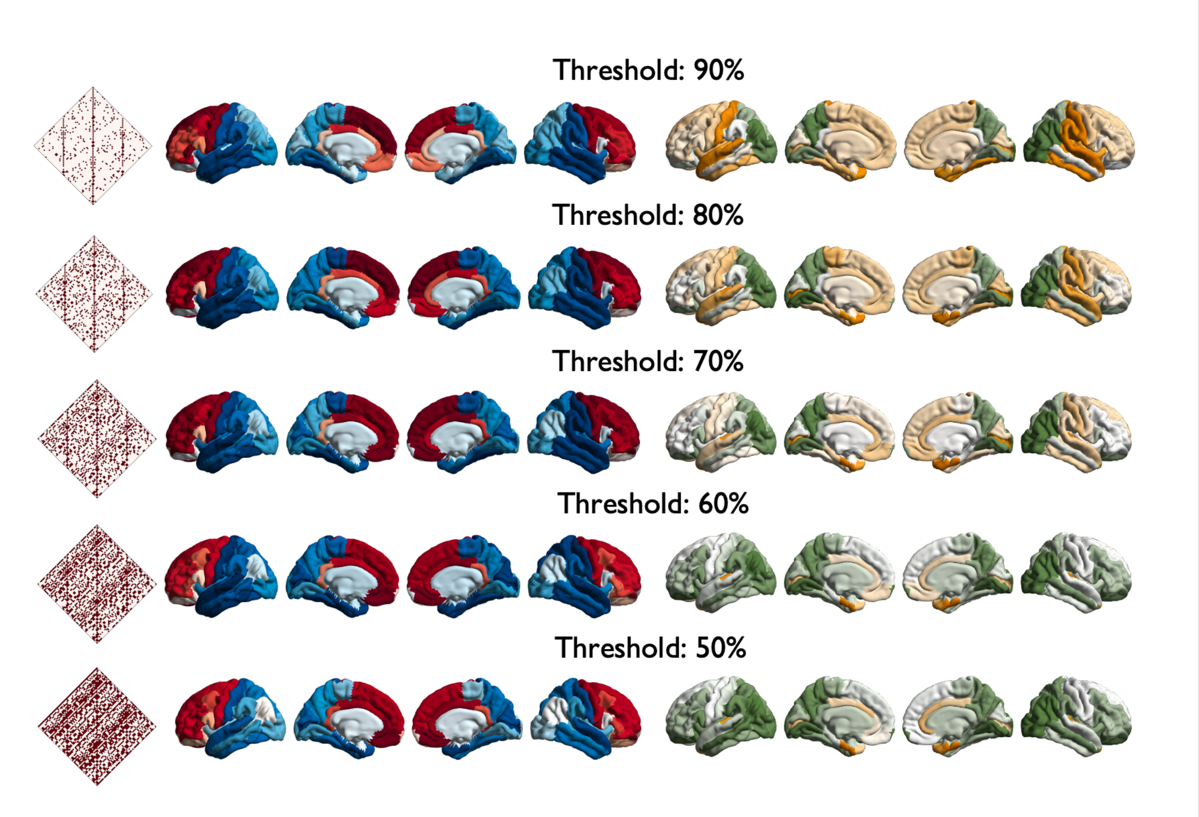


**Figure S2.** *Impact of threshold selection of derived gradients.* We extracted the top 50-90 % of inter-regional cross-disorder correlations for the gradient computation to investigate the impact of threshold selection on the stability of described transdiagnostic gradients.

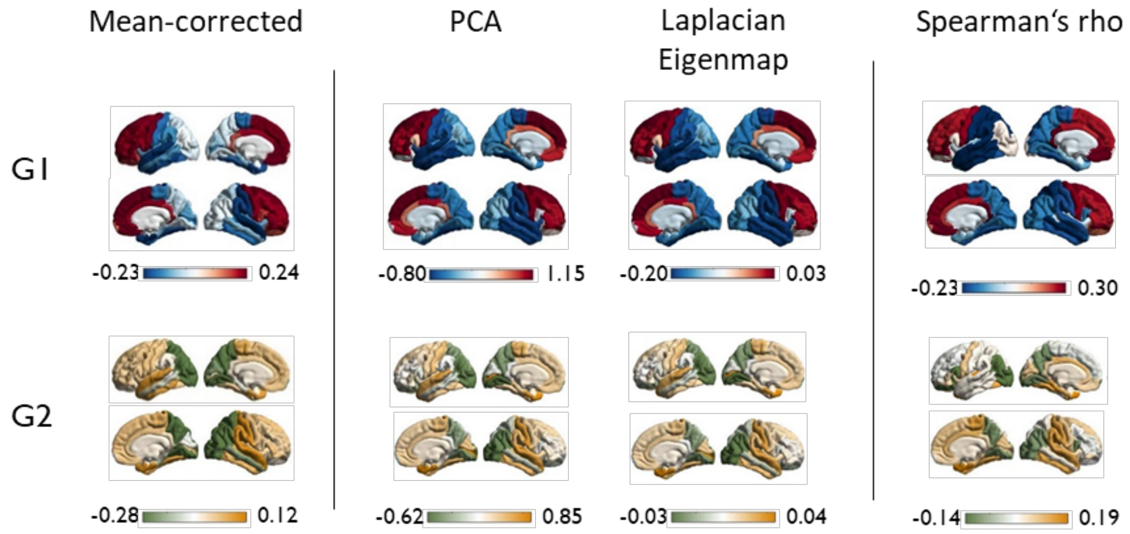


**Figure S3**. *Robustness of transdiagnostic gradients*. We assessed robustness of the two original gradients by comparing them to gradients derived from a mean-corrected correlation matrix (left), gradients based on different dimension reduction methods (middle), and gradients based on a correlation matrix using Spearman's *rho* (right). PCA = principal component analysis.

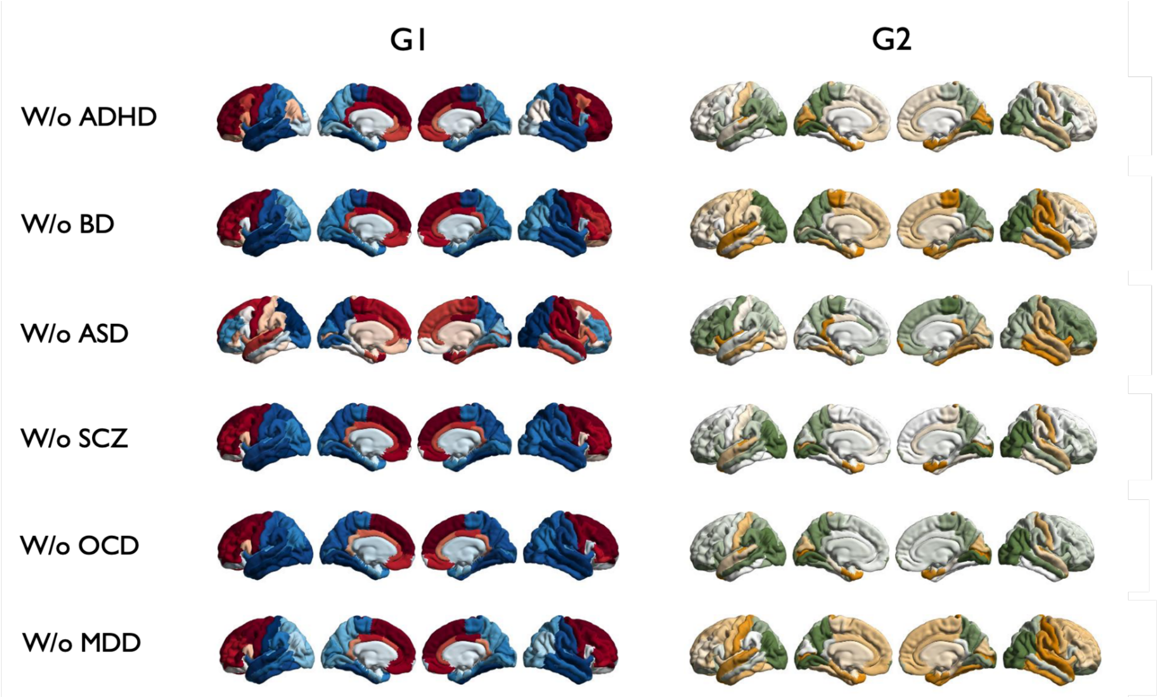


**Figure S4.** *Influence of individual disorders on gradient organization.* We repeated the computation of transdiagnostic gradients using the same parameters as described in the main manuscript (kernel: normalized angel; dimensionality reduction method: diffusion embedding; Cut-off for input correlation matrix: 80%), using different disorder combinations.

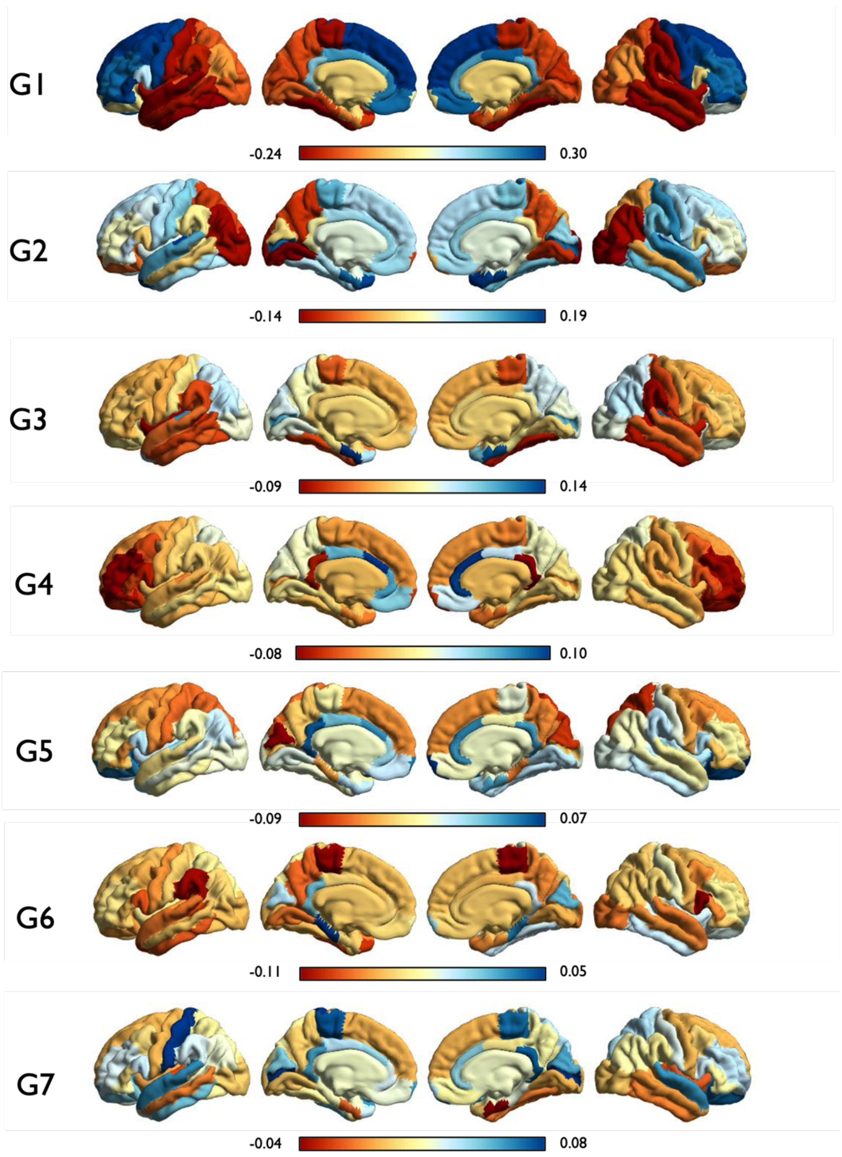


**Figure S5.** *Overview of all gradients computed from cross-disorder correlation matrix using diffusion embedding.*

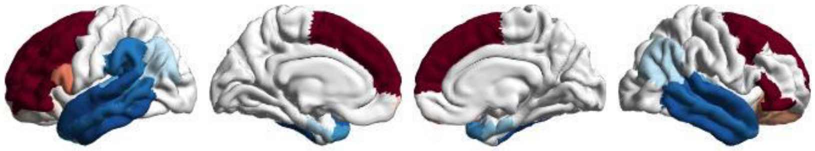

**Figure S6.** *Gradient loadings at epicenters.* Principal axis (G1) masked by significant functional epicenters, demonstrating that epicenters are strongly placed towards apices of the gradient. Red and blue colors indicate opposite apices of G1

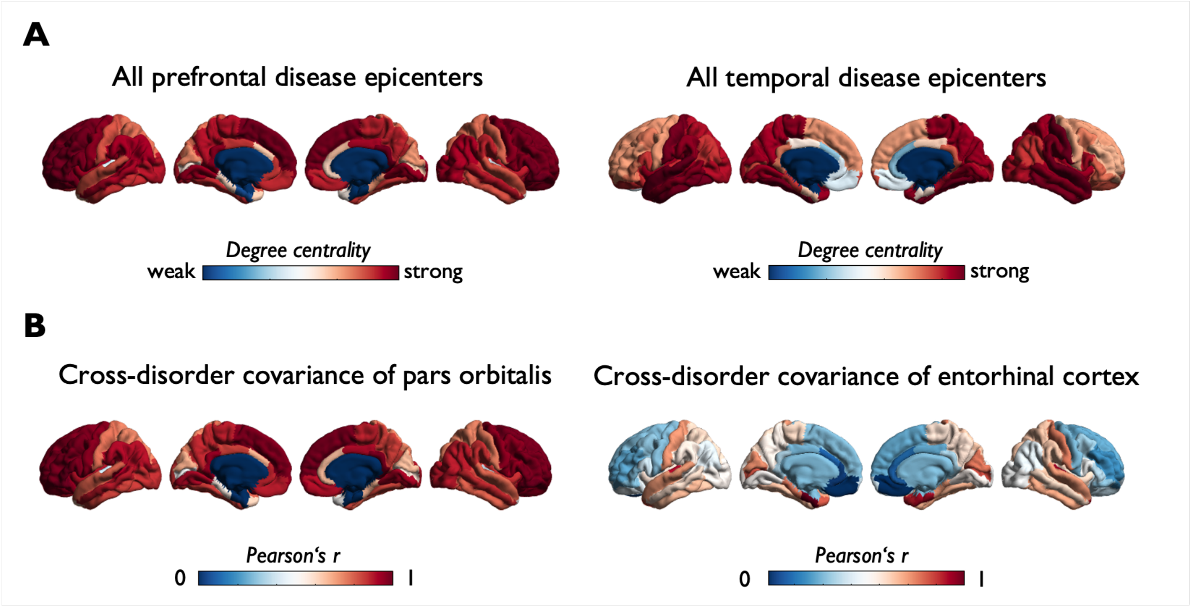


**Figure S7.** *G1 captures segregation of functional disease epicenters.* A) Depicts degree centrality of frontal (left) and temporal (right) functional disease epicenters computed based on whole-brain covariance of cross-disorder disease impact for respective frontal and temporal parcels. B) Shows isolated covariance patterns for the two most likely disease epicenters representative for frontal and temporal structures.

### Supplementary Tables

**Table S1.** Cohen's *d* values describing case-control differences in cortical thickness for 6 mental disorders used for transdiagnostic analyses.

| ROI | ADHD | ASD | BD | MDD | OCD | SCZ |
|---|---|---|---|---|---|---|
| ***L banks sts*** | 0.01 | -0.07 | -0.21 | -0.06 | -0.06 | -0.35 |
| ***L caudal anterior cingulate*** | -0.11 | 0.03 | -0.10 | -0.04 | 0.00 | -0.12 |
| ***L caudal middle frontal*** | -0.02 | 0.06 | -0.27 | -0.01 | -0.09 | -0.36 |
| ***L cuneus*** | 0.11 | -0.06 | -0.06 | 0.05 | -0.04 | -0.20 |
| ***L entorhinal*** | -0.06 | -0.24 | -0.04 | -0.04 | -0.06 | -0.20 |
| ***L fusiform*** | -0.01 | -0.19 | -0.29 | -0.12 | -0.11 | -0.49 |
| ***L inferior parietal*** | 0.08 | -0.05 | -0.27 | -0.06 | -0.14 | -0.36 |
| ***L inferior temporal*** | 0.00 | -0.16 | -0.25 | -0.05 | -0.09 | -0.45 |
| ***L isthmus cingulate*** | 0.04 | 0.05 | -0.13 | -0.10 | -0.07 | -0.31 |
| ***L lateral occipital*** | 0.14 | -0.02 | -0.16 | -0.02 | -0.07 | -0.33 |
| ***L lateral orbitofrontal*** | 0.03 | 0.00 | -0.22 | -0.05 | -0.10 | -0.40 |
| ***L lingual*** | 0.11 | -0.02 | -0.21 | 0.01 | -0.05 | -0.35 |
| ***L medial orbitofrontal*** | -0.08 | 0.08 | -0.20 | -0.13 | -0.08 | -0.23 |
| ***L middle temporal*** | 0.02 | -0.12 | -0.25 | -0.09 | -0.09 | -0.44 |
| ***L parahippocampal*** | 0.10 | -0.11 | -0.02 | -0.07 | -0.06 | -0.28 |
| ***L paracentral*** | -0.01 | -0.05 | -0.14 | 0.00 | -0.01 | -0.25 |
| ***L pars opercularis*** | -0.01 | -0.03 | -0.29 | -0.06 | -0.08 | -0.38 |
| ***L pars orbitalis*** | -0.01 | 0.04 | -0.25 | -0.07 | -0.05 | -0.32 |
| ***L pars triangularis*** | -0.03 | 0.05 | -0.27 | -0.05 | -0.03 | -0.34 |
| ***L pericalcarine*** | 0.04 | -0.01 | 0.02 | 0.09 | 0.01 | -0.08 |
| ***L postcentral*** | 0.05 | -0.07 | -0.10 | 0.04 | -0.02 | -0.26 |
| ***L posterior cingulate*** | -0.11 | 0.05 | -0.11 | -0.10 | -0.07 | -0.30 |
| ***L precentral*** | -0.06 | 0.09 | -0.21 | -0.02 | -0.02 | -0.34 |
| ***L precuneus*** | 0.04 | -0.08 | -0.21 | -0.02 | -0.10 | -0.30 |
| ***L rostral anterior cingulate*** | -0.09 | 0.01 | -0.15 | -0.13 | -0.07 | -0.18 |
| ***L rostral middle frontal*** | 0.03 | 0.11 | -0.28 | -0.04 | -0.10 | -0.36 |
| ***L superior frontal*** | -0.04 | 0.11 | -0.23 | -0.07 | -0.07 | -0.43 |

| | | | | | | |
|---|---|---|---|---|---|---|
| ***L superior parietal*** | 0.10 | -0.09 | -0.16 | -0.01 | -0.06 | -0.21 |
| ***L superior temporal*** | 0.04 | -0.15 | -0.21 | 0.01 | -0.01 | -0.44 |
| ***L supramarginal*** | 0.02 | -0.09 | -0.25 | -0.05 | -0.05 | -0.40 |
| ***L frontal pole*** | 0.08 | 0.04 | -0.12 | -0.01 | -0.05 | -0.21 |
| ***L temporal pole*** | -0.02 | -0.14 | -0.12 | 0.01 | 0.03 | -0.25 |
| ***L transverse temporal*** | -0.02 | -0.24 | -0.12 | -0.04 | 0.00 | -0.25 |
| ***L insula*** | -0.01 | -0.09 | -0.20 | -0.11 | -0.07 | -0.41 |
| ***R bankssts*** | 0.01 | -0.07 | -0.13 | -0.07 | 0.01 | -0.36 |
| ***R caudal anterior cingulate*** | -0.11 | 0.03 | -0.06 | -0.08 | -0.04 | -0.15 |
| ***R caudal middle frontal*** | -0.02 | 0.06 | -0.21 | 0.01 | -0.08 | -0.32 |
| ***R cuneus*** | 0.11 | -0.06 | -0.03 | 0.05 | -0.03 | -0.23 |
| ***R entorhinal*** | -0.06 | -0.24 | -0.08 | -0.06 | 0.01 | -0.15 |
| ***R fusiform*** | -0.01 | -0.19 | -0.27 | -0.12 | -0.09 | -0.54 |
| ***R inferior parietal*** | 0.08 | -0.05 | -0.26 | -0.04 | -0.14 | -0.35 |
| ***R inferior temporal*** | 0.00 | -0.16 | -0.19 | -0.12 | -0.06 | -0.44 |
| ***R isthmus cingulate*** | 0.04 | 0.05 | -0.18 | -0.07 | -0.05 | -0.31 |
| ***R lateral occipital*** | 0.14 | -0.02 | -0.22 | 0.01 | -0.07 | -0.34 |
| ***R lateral orbitofrontal*** | 0.03 | 0.00 | -0.21 | -0.12 | -0.11 | -0.36 |
| ***R lingual*** | 0.11 | -0.02 | -0.20 | -0.01 | -0.04 | -0.39 |
| ***R medial orbitofrontal*** | -0.08 | 0.08 | -0.23 | -0.13 | -0.10 | -0.24 |
| ***R middle temporal*** | 0.02 | -0.12 | -0.22 | -0.09 | -0.10 | -0.38 |
| ***R parahippocampal*** | 0.10 | -0.11 | -0.09 | -0.06 | -0.08 | -0.29 |
| ***R paracentral*** | -0.01 | -0.05 | -0.14 | -0.01 | 0.01 | -0.22 |
| ***R pars opercularis*** | -0.01 | -0.03 | -0.25 | -0.02 | -0.06 | -0.42 |
| ***R pars orbitalis*** | -0.01 | 0.04 | -0.24 | -0.07 | -0.07 | -0.34 |
| ***R pars triangularis*** | -0.03 | 0.05 | -0.23 | -0.03 | -0.06 | -0.37 |
| ***R pericalcarine*** | 0.04 | -0.01 | 0.02 | 0.08 | 0.03 | -0.09 |
| ***R postcentral*** | 0.05 | -0.07 | -0.08 | 0.03 | 0.04 | -0.28 |
| ***R posterior cingulate*** | -0.11 | 0.05 | -0.17 | -0.09 | -0.06 | -0.31 |
| ***R precentral*** | -0.06 | 0.09 | -0.18 | -0.02 | -0.04 | -0.32 |
| ***R precuneus*** | 0.04 | -0.08 | -0.19 | 0.01 | -0.10 | -0.30 |
| ***R rostral anterior cingulate*** | -0.09 | 0.01 | -0.09 | -0.10 | 0.01 | -0.12 |

| | | | | | | |
|---|---|---|---|---|---|---|
| ***R rostral middle frontal*** | 0.03 | 0.11 | -0.26 | -0.04 | -0.09 | -0.31 |
| ***R superior frontal*** | -0.04 | 0.11 | -0.26 | -0.08 | -0.04 | -0.40 |
| ***R superior parietal*** | 0.10 | -0.09 | -0.16 | 0.03 | -0.05 | -0.22 |
| ***R superior temporal*** | 0.04 | -0.15 | -0.19 | -0.03 | 0.01 | -0.44 |
| ***R supramarginal*** | 0.02 | -0.09 | -0.18 | -0.05 | 0.00 | -0.39 |
| ***R frontal pole*** | 0.08 | 0.04 | -0.10 | -0.06 | 0.02 | -0.21 |
| ***R temporal pole*** | -0.02 | -0.14 | -0.06 | 0.01 | 0.02 | -0.24 |
| ***R transverse temporal*** | -0.02 | -0.24 | -0.11 | -0.05 | -0.02 | -0.26 |
| ***R insula*** | -0.01 | -0.09 | -0.17 | -0.12 | -0.07 | -0.41 |

Cohen's *d* values were accessed through the ENIGMA Toolbox (3) and for adult samples (except for Autism spectrum disorder (ASD), for which data was only available for a pooled sample including younger subjects). Data was collected and analyzed by respective ENIGMA working groups (Attention-deficit/hyperactivity disorder (ADHD) (6), Autism spectrum disorder (ASD) (7), Bipolar disorder (BD) (8), Major depressive disorder (MDD) (9), Obsessive compulsive disorder (OCD) (10), Schizophrenia (SCZ) (11)) and
adjusted for different combinations of age, sex, scan site/scanner differences, intracranial volume, and intelligence quotient effects (see Supplementary Table 4).

**Table S2. Sample Demographics.**

| Disorder | sites | Weighted mean age (cases) | Weighted mean age (controls) | *n* | Covariates |
|---|---|---|---|---|---|
| **Schizophrenia (11)** | 39 | 32.3[a] | 34.5[a] | Cases: 4474<br>Controls: 5098<br>Total: 9572 | age, sex, scan site |
| **Attention deficit hyperactivity disorder (6)** | 36 | 32.97 | | Cases: 733<br>Controls: 539<br>Total: 1272 | age, sex |
| **Autism spectrum disorder (7)** | 49 | 15.4 years | 15.8 years | Cases: 1571<br>Controls: 1651<br>Total: 3222 | age, sex, IQ |
| **Bipolar Disorder (8)** | 28 | 38.4[a] | 35.6[a] | Cases: 1837<br>Controls: 2582<br>Total: 4419 | age, sex |
| **Major depressive disorder (9)** | 20 | 44.8[a] | 54.6[a] | Cases: 1911<br>Controls: 7663<br>Total: 9574 | age, sex, scan site |
| **Obsessive-compulsive disorder (10)** | 27 | 32.1 | 30.5 | Cases: 1,498<br>Controls: 1,436<br>Total: 2934 | age, sex, scan site |

Adapted from Radonjić et al. (12). [a] = weighted mean computed by Radonjić et al. (12). IQ = Intelligence quotient.

**Table S3.** Link between principal (G1) and secondary (G2) transdiagnostic axes of pathological covariance and disease-specific Cohen's *d* maps.

| | **ADHD** | **ASD** | **BD** | **MDD** | **SCZ** | **OCD** |
|---|---|---|---|---|---|---|
| **G1** | $r$ = -0.52, $p_{spin}$ = 0.002 | $r$ = 0.82, $p_{spin}$ = 0.001 | n.s. | n.s. | n.s. | $r$ = -0.20, $p_{spin}$ = 0.021 |
| **G2** | $r$ = -0.58, $p_{spin}$ < .0001 | $r$ = -0.26, $p_{spin}$ = 0.050 | $r$ = 0.42, $p_{spin}$ = 0.001 | n.s. | $r$ = 0.24, $p_{spin}$ = 0.041 | $r$ = 0.6, $p_{spin}$ = 0.001 |

**Table S4.** 199 Genes for which spatial transcription patterns correlated significantly with the principal transdiagnostic gradient.

| Genes | *r* | p | Genes | *r* | p | Genes | *r* | p |
|---|---|---|---|---|---|---|---|---|
| **PRRX1** | 0.8 | 6.05E-02 | **EPHB3** | 0.55 | 1.77E+08 | **CENPJ** | -0.5 | 7.61E+08 |
| **ZIC1** | 0.74 | 9.23E+01 | **COL11A1** | 0.55 | 1.81E+08 | **VCX** | -0.5 | 7.02E+08 |
| **CTXN3** | 0.71 | 2.47E+03 | **LINC02217** | 0.55 | 1.89E+08 | **PLK5** | -0.5 | 6.90E+08 |
| **RPH3AL** | 0.7 | 6.88E+03 | **HRH3** | 0.55 | 1.94E+08 | **RASIP1** | -0.5 | 6.78E+08 |
| **CD6** | 0.69 | 1.56E+04 | **SKAP1** | 0.55 | 1.96E+08 | **ANKRD20A11P** | -0.5 | 6.76E+08 |
| **LAMA2** | 0.68 | 2.32E+04 | **LCP2** | 0.55 | 2.09E+08 | **SPIDR** | -0.5 | 6.68E+08 |
| **HSPB8** | 0.67 | 5.28E+04 | **GPR26** | 0.55 | 2.18E+08 | **DUSP8** | -0.5 | 5.00E+08 |
| **KRT31** | 0.67 | 5.83E+04 | **MEIS3P1** | 0.55 | 2.22E+08 | **PATJ** | -0.5 | 4.78E+08 |
| **WNT10A** | 0.67 | 9.27E+04 | **ASL** | 0.54 | 2.24E+08 | **ADAMTSL1*** | -0.5 | 4.36E+08 |
| **ACTC1** | 0.67 | 9.50E+04 | **NUDT14** | 0.54 | 2.25E+08 | **BOLA2_SMG1P6** | -0.5 | 4.33E+08 |
| **YBX2** | 0.66 | 1.21E+05 | **TMTC3** | 0.54 | 2.39E+08 | **GLIS1** | -0.5 | 4.09E+08 |
| **GORAB** | 0.65 | 2.49E+05 | **EFHC2*** | 0.54 | 2.53E+08 | **MTHFD2L** | -0.5 | 3.64E+08 |
| **FOXF2*** | 0.65 | 3.26E+05 | **ALPL*** | 0.54 | 2.54E+08 | **NME5** | -0.5 | 3.45E+08 |
| **CCDC80** | 0.64 | 7.77E+05 | **TGFBI** | 0.54 | 2.87E+08 | **TRANK1*** | -0.5 | 3.36E+08 |
| **CD38** | 0.64 | 8.09E+05 | **CCDC110** | 0.54 | 3.05E+08 | **FBXO11** | -0.5 | 3.15E+08 |
| **TCERG1L** | 0.64 | 8.16E+05 | **HGF** | 0.54 | 3.17E+08 | **COL5A1** | -0.5 | 3.02E+08 |
| **TEX30** | 0.64 | 8.66E+05 | **VAT1** | 0.54 | 3.32E+08 | **MIS18BP1** | -0.5 | 2.81E+08 |
| **NRP1** | 0.64 | 1.02E+06 | **CMTM4** | 0.54 | 3.44E+08 | **PLOD2** | -0.5 | 2.60E+08 |

| | | | | | | | | |
|---|---|---|---|---|---|---|---|---|
| **CREB3L3** | 0.63 | 1.77E+06 | **NANOS3** | 0.54 | 3.50E+08 | **RIPK1** | -0.5 | 2.41E+08 |
| **SPRY4** | 0.63 | 1.80E+06 | **CHST8** | 0.54 | 3.63E+08 | **DPP6** | -0.5 | 2.39E+08 |
| **CBLN2** | 0.62 | 2.26E+06 | **SIX3** | 0.53 | 3.91E+08 | **TENM2*** | -0.5 | 2.24E+08 |
| **PCDH10** | 0.62 | 2.33E+06 | **ASB6** | 0.53 | 4.41E+08 | **CRIP3** | -0.5 | 2.09E+08 |
| **FAM213A** | 0.62 | 2.73E+06 | **MELTF** | 0.53 | 4.57E+08 | **HYDIN2** | -0.5 | 1.90E+08 |
| **SULF1** | 0.62 | 2.76E+06 | **LOC100128239** | 0.53 | 4.59E+08 | **WNT3** | -0.6 | 1.60E+08 |
| **HDAC9** | 0.62 | 3.42E+06 | **ASB2** | 0.53 | 4.60E+08 | **GREB1L** | -0.6 | 1.52E+08 |
| **ARHGAP25*** | 0.61 | 4.31E+06 | **LBH** | 0.53 | 5.04E+08 | **SDHAF4** | -0.6 | 1.04E+08 |
| **RTP1** | 0.6 | 7.89E+06 | **ALKBH7*** | 0.53 | 5.07E+08 | **MST1R** | -0.6 | 9.88E+07 |
| **AEBP1** | 0.6 | 1.07E+07 | **FLJ30901** | 0.53 | 5.21E+08 | **PDGFD** | -0.6 | 8.93E+07 |
| **TSPAN33** | 0.6 | 1.09E+07 | **SLC35F1** | 0.53 | 5.23E+08 | **DNAAF5** | -0.6 | 7.91E+07 |
| **PROCA1** | 0.6 | 1.36E+07 | **TRAF3*** | 0.53 | 5.42E+08 | **TENM4** | -0.6 | 7.29E+07 |
| **AMDHD1** | 0.6 | 1.37E+07 | **KCNA5** | 0.52 | 6.77E+08 | **THSD7A** | -0.6 | 5.25E+07 |
| **TNFRSF14** | 0.59 | 1.50E+07 | **KLF6** | 0.52 | 6.83E+08 | **MET** | -0.6 | 3.66E+07 |
| **LGALS1** | 0.59 | 1.51E+07 | **SPON2** | 0.52 | 7.08E+08 | **SLFN11** | -0.6 | 3.58E+07 |
| **RBP1** | 0.59 | 1.54E+07 | **SYNM** | 0.52 | 7.16E+08 | **LAMP5** | -0.6 | 3.52E+07 |
| **WFDC1** | 0.59 | 2.35E+07 | **FDPS** | 0.52 | 7.24E+08 | **ZNF662** | -0.6 | 3.52E+07 |
| **GALNT16** | 0.59 | 2.47E+07 | **PLCH1** | 0.52 | 7.62E+08 | **SYN3** | -0.6 | 3.36E+07 |
| **AZIN2** | 0.58 | 2.70E+07 | **ADAMTS9** | 0.52 | 7.86E+08 | **TPTE2P1** | -0.6 | 2.02E+07 |

| | | | | | | | | |
|---|---|---|---|---|---|---|---|---|
| **MRAP2** | 0.58 | 2.78E+07 | **DZIP3** | 0.52 | 7.98E+08 | **MGP** | -0.6 | 9.92E+06 |
| **CDH13*** | 0.58 | 2.79E+07 | **CLIC5** | 0.52 | 8.24E+08 | **IL17RA** | -0.6 | 9.80E+06 |
| **DBX2** | 0.58 | 2.83E+07 | **SLC35F2*** | 0.52 | 8.50E+08 | **TMEM249** | -0.6 | 7.31E+06 |
| **ERICH1** | 0.58 | 2.92E+07 | **GPX3** | 0.52 | 9.18E+08 | **CDH12** | -0.6 | 5.24E+06 |
| **OVOL2** | 0.58 | 3.18E+07 | **PEBP4** | 0.52 | 9.31E+08 | **PAX8_AS1** | -0.6 | 3.22E+06 |
| **ABHD12B*** | 0.58 | 3.22E+07 | **DDAH1** | 0.52 | 9.45E+08 | **DSP** | -0.6 | 2.37E+06 |
| **ZC2HC1A** | 0.58 | 3.32E+07 | **STK17A** | 0.52 | 9.46E+08 | **ARL9** | -0.6 | 6.32E+05 |
| **SCRG1** | 0.58 | 3.64E+07 | **CNTN6** | 0.51 | 9.68E+08 | **SLC17A6** | -0.7 | 1.83E+05 |
| **ITGA8** | 0.58 | 3.68E+07 | **ADTRP** | 0.51 | 1.02E+09 | **CPLX2** | -0.7 | 8.40E+04 |
| **KCNN2** | 0.58 | 4.02E+07 | **RARA_AS1** | 0.51 | 1.05E+09 | **ANKRD20A5P** | -0.7 | 2.15E+04 |
| **NT5DC2*** | 0.58 | 4.05E+07 | **TSTA3** | 0.51 | 1.10E+09 | **C15orf59** | -0.7 | 1.31E+04 |
| **HSPB3** | 0.57 | 4.88E+07 | **CDT1** | 0.51 | 1.15E+09 | **COL27A1** | -0.7 | 3.37E+03 |
| **CYP51A1** | 0.57 | 5.17E+07 | **MSRB2** | 0.51 | 1.16E+09 | **WDR97** | -0.7 | 2.56E+03 |
| **TEX26** | 0.57 | 5.40E+07 | **CYP26A1** | 0.51 | 1.16E+09 | **LXN** | -0.7 | 2.41E+02 |
| **TMEM117** | 0.57 | 5.72E+07 | **S1PR2** | 0.51 | 1.24E+09 | | | |
| **ASGR2** | 0.57 | 6.50E+07 | **CASC10** | 0.51 | 1.26E+09 | | | |
| **C6orf62** | 0.57 | 6.96E+07 | **WNT7A** | 0.51 | 1.27E+09 | | | |
| **CIB1** | 0.57 | 7.31E+07 | **THBD** | 0.51 | 1.28E+09 | | | |
| **CTSH** | 0.57 | 7.57E+07 | **PERM1** | 0.51 | 1.29E+09 | | | |
| **GAL** | 0.56 | 8.34E+07 | **GATB** | 0.51 | 1.30E+09 | | | |
| **FSTL5** | 0.56 | 9.28E+07 | **BCL7C** | 0.51 | 1.41E+09 | | | |
| **HAUS7** | 0.56 | 9.37E+07 | **BCKDK** | 0.51 | 1.43E+09 | | | |
| **NR4A3** | 0.56 | 1.08E+08 | **BNIP3** | 0.51 | 1.48E+09 | | | |
| **ATP2C2** | 0.56 | 1.15E+08 | **MGAT4C*** | 0.51 | 1.51E+09 | | | |

| | | | | | |
|---|---|---|---|---|---|
| **CHRNA6** | 0.56 | 1.21E+08 | **FABP6** | 0.51 | 1.53E+09 |
| **C10orf82** | 0.56 | 1.22E+08 | **AKAP13*** | -0.5 | 1.44E+09 |
| **BAIAP2L 2** | 0.55 | 1.36E+08 | **TIAM1** | -0.5 | 1.38E+09 |
| **TNC** | 0.55 | 1.38E+08 | **EPB41L1** | -0.5 | 1.38E+09 |
| **RAB3C** | 0.55 | 1.41E+08 | **ACAP3** | -0.5 | 1.36E+09 |
| **VIT** | 0.55 | 1.41E+08 | **PPFIA4** | -0.5 | 1.20E+09 |
| **NECTIN3** | 0.55 | 1.48E+08 | **ZNF785** | -0.5 | 1.15E+09 |
| **MND1** | 0.55 | 1.53E+08 | **BTBD3** | -0.5 | 1.13E+09 |
| **TUNAR** | 0.55 | 1.55E+08 | **RCC1L** | -0.5 | 1.05E+09 |
| **ZIC3** | 0.55 | 1.57E+08 | **KRTCAP3** | -0.5 | 9.91E+08 |
| **RIT2** | 0.55 | 1.65E+08 | **PAM** | -0.5 | 8.66E+08 |
| **PPEF1** | 0.55 | 1.71E+08 | **NPR3** | -0.5 | 7.77E+08 |
| **GPR143** | 0.55 | 1.73E+08 | **IGDCC3*** | -0.5 | 7.68E+08 |

*Gene has been identified to be a risk gene for one of the included disorders in recent genome-wide association studies. Genes linked to attention-deficit hyperactivity disorder: ABHD12B and ALKBH7 (13); Autism spectrum disorder: ALPL (14); Bipolar disorder: ADAMTSL1, AKAP13, and ARHGAP25 (15); Major depressive disorder: CDH13, EFHC2, IGDCC3, MGAT4C, TENM2, and TRAF3 (16); Schizophrenia: FOXF2, NT5DC2, SLC35F2, and TRANK1 (17).
